# Dose-dependent volume loss in subcortical deep grey matter structures after cranial radiotherapy

**DOI:** 10.1101/2020.07.23.20160606

**Authors:** Steven H.J Nagtegaal, Szabolcs David, Marielle E.P. Philippens, Tom J. Snijders, Alexander Leemans, Joost J.C. Verhoeff

**Affiliations:** Department of Radiation Oncology, University Medical Center Utrecht, The Netherlands, HP Q 00.3.11, PO box 85500, 3508 GA, Utrecht, the Netherlands; UMC Utrecht Brain Center, Department of Neurology & Neurosurgery, University Medical Center Utrecht, The Netherlands, HP L 01.310, PO box 85500, 3508 GA, Utrecht, the Netherlands; Image Sciences Institute, University Medical Center Utrecht, The Netherlands, HP Q 00.3.11, PO box 85500, 3508 GA, Utrecht, the Netherlands

**Keywords:** Radiotherapy, Brain Neoplasms, Gray Matter, Amygdala, Nucleus accumbens, Caudate nucleus, Hippocampus, Globus pallidus, Putamen, Thalamus

## Abstract

**Background and purpose:** The relation between radiotherapy (RT) dose to the brain and morphological changes in healthy tissue has seen recent increased interest. There already is evidence for changes in the cerebral cortex and white matter, as well as selected subcortical grey matter (GM) structures. We studied this relation in all deep GM structures, to help understand the aetiology of post-RT neurocognitive symptoms.

**Materials and methods:** We selected 31 patients treated with RT for glioma. Pre-RT and post-RT 3D T1 MRIs were automatically segmented, and the changes in volume of the following structures were assessed: amygdala, nucleus accumbens, caudate nucleus, hippocampus, globus pallidus, putamen, and thalamus. The volumetric changes were related to the mean RT dose received by each structure. Hippocampal volumes were entered into a population-based nomogram to estimate hippocampal age.

**Results:** A significant relation between RT dose and volume loss was seen in all examined structures, except the caudate nucleus. The volume loss rates ranged from 0.16-1.37 %/Gy, corresponding to 4.9-41.2% per 30 Gy. Hippocampal age, as derived from the nomogram, was seen to increase by a median of 11 years.

**Conclusion:** Almost all subcortical GM structures are susceptible to radiation-induced volume loss, with more volume loss being observed with increasing dose. Volume loss of these structures is associated with neurological deterioration, including cognitive decline, in neurodegenerative diseases. To support a causal relationship between radiation-induced deep GM loss and neurocognitive functioning in glioma patients, future studies are needed that directly correlate volumetrics to clinical outcomes.

## Introduction

Irradiation of healthy brain tissue can lead to anatomical and functional deficits, a phenomenon known as radiation-induced brain injury. This can lead to a variety of symptoms, with especially cognitive and executional impairments leading to a marked decrease in the patient’s quality of life after radiation therapy (RT) [1,2].

With the advent of high-resolution brain imaging, the interest in morphological changes after RT has increased. The cerebral cortex has been shown to be susceptible to radiation-induced thinning, especially in areas associated with cognitive functioning [3–6]. Thinning rates are found to be dose-dependent, meaning that a higher dose leads to a further diminished cortex. Similarly, diffusion tensor imaging has shown that white matter shows dose-dependent changes in several metrics after RT [7]. Finally, two grey matter structures, the hippocampus [8] and the amygdala [9], show susceptibility to radiation damage, again with higher volume changes with increasing dose. Furthermore, the dose to the hippocampus has been shown to negatively affect neurocognitive outcome after RT [10].

Less is known about the susceptibility to radiation damage of other subcortical grey matter structures, such as the nucleus accumbens, caudate nucleus, globus pallidus, putamen, and thalamus. Atrophy of these deep GM structures is associated with impaired cognitive function in patients with degenerative brain diseases as well as healthy ageing [11–13]. This relation is most pronounced in Alzheimer’s disease, with the volume of all mentioned structures, with the exception of globus pallidus, being associated with cognitive impairment [14–16]. Globus pallidus volume in its turn is associated with cognitive outcomes in Huntington’s disease and age-related cognitive impairments [17,18].

Volume changes in these structures are associated with cognitive outcomes, and the cause of post-RT cognitive decline needs to be elucidated. Therefore, we examined the relation between post-RT subcortical GM volume changes and RT dose.

## Methods

### Patient selection and data collection

Patients who were treated with RT for newly discovered grade II-IV glioma at the department of Radiation Oncology in 2016 and 2017 were retrospectively identified. Criteria for inclusion were: treatment planning CT and MRI present, with isotropic high resolution; survival > 270 days after RT; and availability of at least 1 follow-up MRI between 270 days and 360 days after RT, and with isotropic high resolution. Clinical MRI and CT scans made for RT treatment planning, all follow-up MRIs, and clinical and demographic characteristics were extracted from patient records. The need for informed consent for this retrospective study was waived by our institutional review board (#18/274).

### Image acquisition

For every patient the pre-RT CT and MRI were collected, as well as all available follow-up MRIs. RT planning CT scans were acquired on a Brilliance Big bore scanner (Philips Medical Systems, Best, The Netherlands), with a tube potential of 120 kVp, with a matrix size of 512 × 512 and 0.65 × 0.65 × 3.0 mm voxel size. MR images were acquired on a 3T Philips Ingenia scanner (Philips Healthcare, Best, The Netherlands) as part of routine clinical care. T1-weighted MR images were acquired with a 3D turbo field echo (TFE) sequence without gadolinium enhancement with the following parameters: TR = 8.1 ms, TE = 3.7 ms, flip angle = 8°, matrix: 207 × 289 × 213, and a reconstructed voxel resolution of 1×0.96×0.96 mm.

### Image processing

A graphical overview of the image processing pipeline is shown in **Figure 1**. All imaging data was processed with Statistical Parametric Mapping (SPM12, v7487)[19] Computational Anatomy Toolbox (CAT12.6 r1450) [20] and in-house algorithms developed in MATLAB (Mathworks, Natick, Massachusetts, USA).

**Figure 1.**
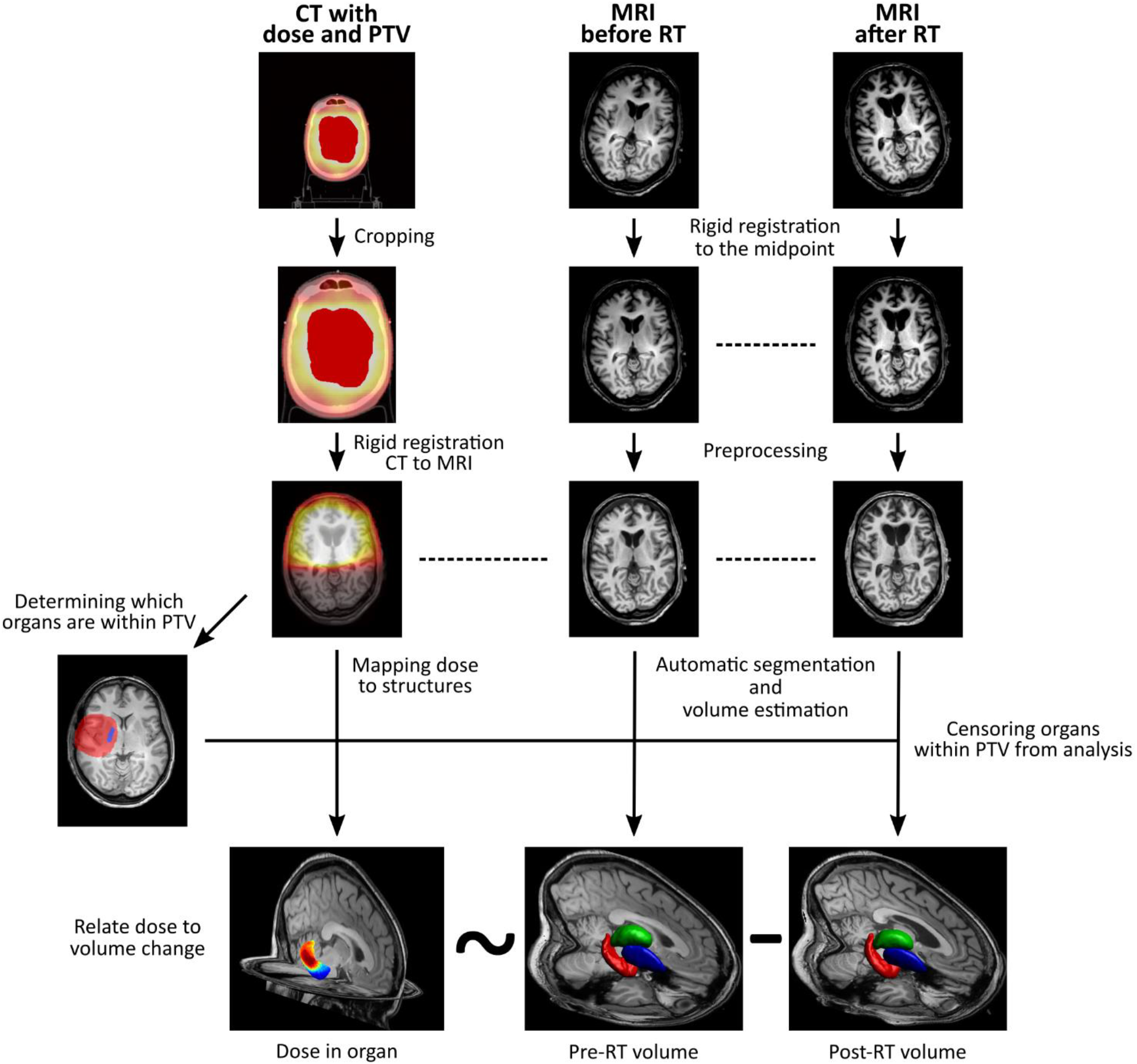
Pipeline of image processing. Left column: dose (gradient) and PTV (red shading) are extracted from CT. Middle and right column: organ volume is estimated from processed MRI. After censoring organs within PTV, applied dose is related to the change in organ volume.

Image processing was done in concordance to our own previously published criteria [3], amended for the current research question. More detailed image processing methods can be found in our previous work [4].

In brief, the cropped CT image with the associated dose and planning target volume (PTV) maps were registered to the T1 MR images, resulting in the CT image and the MRIs being in the same space. Next, the rigidly coregistered T1s were processed with CAT12’s segmentation pipeline.

Deep GM structure volumes were estimated with CAT12 using the fully automated volume estimation method using the labels from the Neuromorphometrics atlas (Neuromorphometrics Inc., Somerville, Massachusetts, USA). The following structures were examined: amygdala, nucleus accumbens, caudate nucleus, hippocampus, globus pallidus, putamen, and thalamus. **Figure 2** shows the anatomical location of the structures on axial T1 MRI, as well as in a 3D rendering.

**Figure 2.**
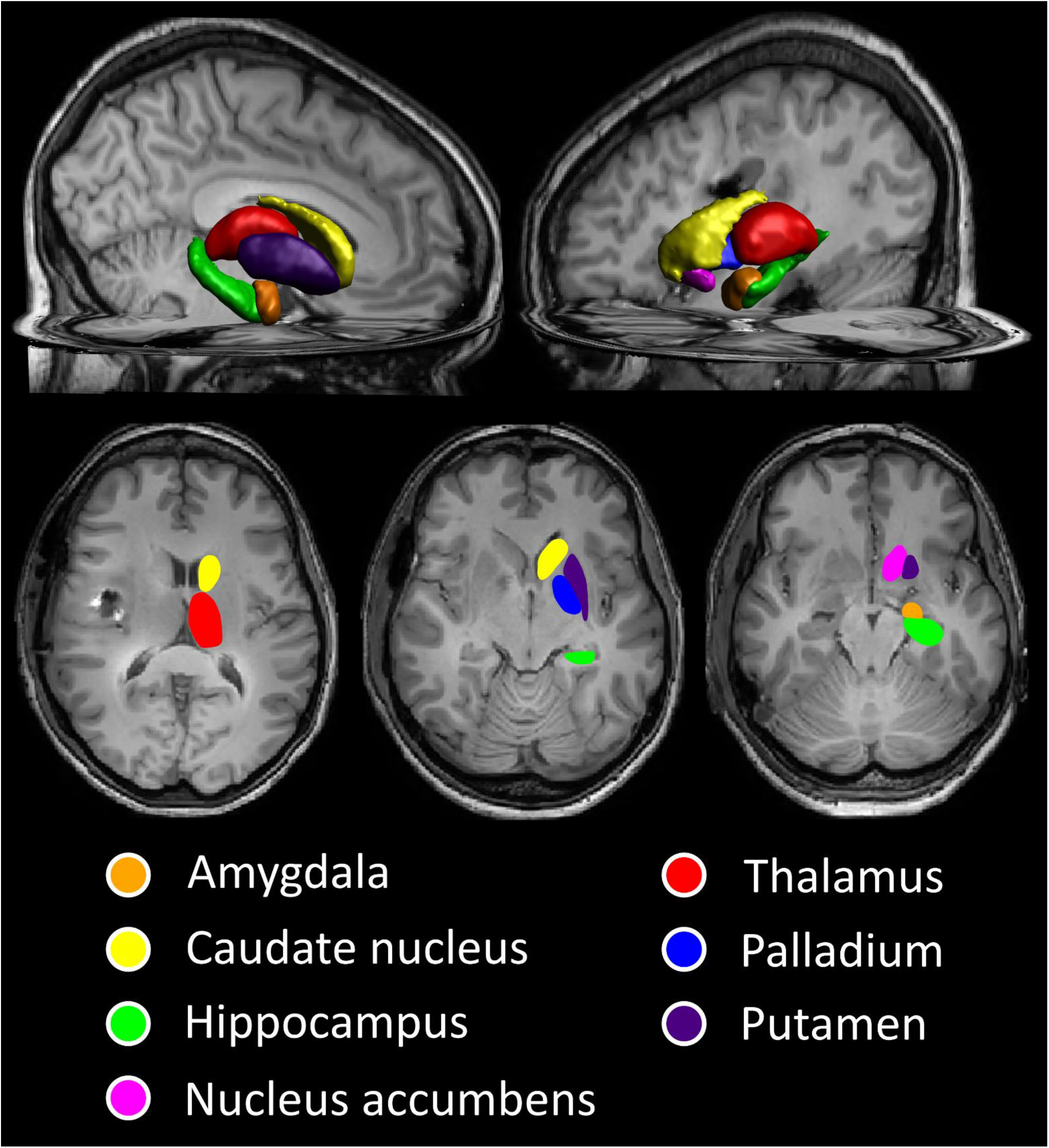
A 3D rendering and axial MR images showing the subcortical grey matter structures being analysed.

The within-subject difference in deep GM volume was calculated by subtracting the baseline and the follow-up volume. In every subject the deep GM organs included in the PTV were censored from analysis, to avoid spurious volume-dose relations originating from segmentation errors due to damage around the tumour. If the residual damage (e.g. oedema, surgical scarring, tumour bed) extended beyond the PTV in either the baseline or the follow-up images, then the affected subject was removed from the analysis.

### Statistical analysis

Statistical comparison of deep GM volume change and dose correlation was carried out with a permutation test with 10,000 iterations performed with the permutation analysis of linear models (PALM) toolbox in Matlab.[21–23] Significance of a correlation was set at pcorr < 0.05, with use of family-wise error rate (FWER) adjustment to correct for multiple comparisons. All further presented p-values are FWER-corrected. Age at the time of the diagnosis and sex of the patients were included as nuisance regressors.

To assess whether administration of chemotherapy has an effect on the relation between dose and volume, a sensitivity analysis was performed in which chemotherapy was added as a covariate to the permutation test, again with FWER-adjustment.

### Hippocampal nomograms

To put volumetric changes of the hippocampus into context, the pre-RT and post-RT volumes were entered into a nomogram of hippocampal volume across age groups. [24,25] This nomogram is based on MRI data from 19,700 healthy participants from the UK Biobank. We did this in two ways: 1) the patients’ new “hippocampal age” was determined based on its volume after RT and compared to the actual age, and 2) we assessed whether there was a change in a patient’s hippocampal volume percentile within the population between the pre-RT and post-RT scans. Due to the age range of the nomograms, only patients aged 52 to 72 could be entered into the nomogram.

## Results

### Participants

Of all the patients treated with RT for glioma in 2016 and 2017, thirty-one patients were eligible for inclusion in the current analysis. A flow-chart of study inclusion in shown in **Supplementary Figure 1**. Extensive damage outside the censored PTV area on baseline MRI meant exclusion of one case. Baseline characteristics are shown in **Table 1**.

**Table 1.** Baseline characteristics of included patients

|  | **N (total n = 31)** |
| --- | --- |
| **Age (mean; SD)** | 50 ($\pm 15$) |
| **Sex** |  |
| Male | 19 (61.3%) |
| Female | 12 (38.7%) |
| **WHO grade** |  |
| II | 12 (38.7%) |
| III | 6 (19.4%) |
| IV | 13 (41.9%) |
| **Tumour type** |  |
| Astrocytoma, IDH-mutant | 13 (41.9%) |
| Astrocytoma, IDH-wildtype | 3 (9.6%) |
| Glioblastoma, IDH-wildtype | 9 (29.0%) |
| Other | 6 (19.6%) |
| **Prescribed dose** |  |
| 28 × 1.8Gy = 50.4 Gy | 11 (35.5%) |
| 30 × 1.8Gy = 54 Gy | 2 (6.5%) |
| 30 × 2.0Gy = 60 Gy | 18 (58.1%) |
| **Concurrent or sequential systemic therapy** |  |
| None | 5 (16.1%) |
| Temozolomide | 21 (67.7%) |
| PCV | 4 (12.9%) |
| PCV = procarbazine, lomustine and vincristine |  |

### Subcortical GM volume

Significant dose-dependent volume loss was observed in all examined structures, except for caudate nucleus. Rates of volume loss vary from 0.16 to 1.37% per Gy (corresponding to 4.9% and 41.2% per 30 Gy), and are shown for all structures in **Table 2**. Scatterplots of the organs in which a significant relation between RT dose and volume loss was seen are shown in **Figure 3**.

**Table 2.** Dose-dependent changes in volumes of subcortical GM structures

|  | n | Volume loss rate (%/Gy) | Volume loss rate (%/30Gy) | 95% confidence interval | p* |
| --- | --- | --- | --- | --- | --- |
| Amygdala | 48 | 0.30 | 9.0 | 0.15 – 0.45 | **<0.01** |
| Caudate nucleus | 38 | 0.10 | 3.0 | -0.22 – 0.42 | 0.74 |
| Globus pallidus | 35 | 1.37 | 41.2 | 0.68 – 2.07 | **<0.01** |
| Hippocampus | 42 | 0.16 | 4.9 | 0.02 – 0.31 | **0.03** |
| Nucleus accumbens | 45 | 0.32 | 9.5 | 0.13 – 0.50 | **<0.01** |
| Putamen | 44 | 0.81 | 24.3 | 0.42 – 1.20 | **<0.01** |
| Thalamus | 29 | 1.15 | 34.5 | 0.74 – 1.56 | **<0.01** |

\*corrected for multiple testing

**Figure 3.**
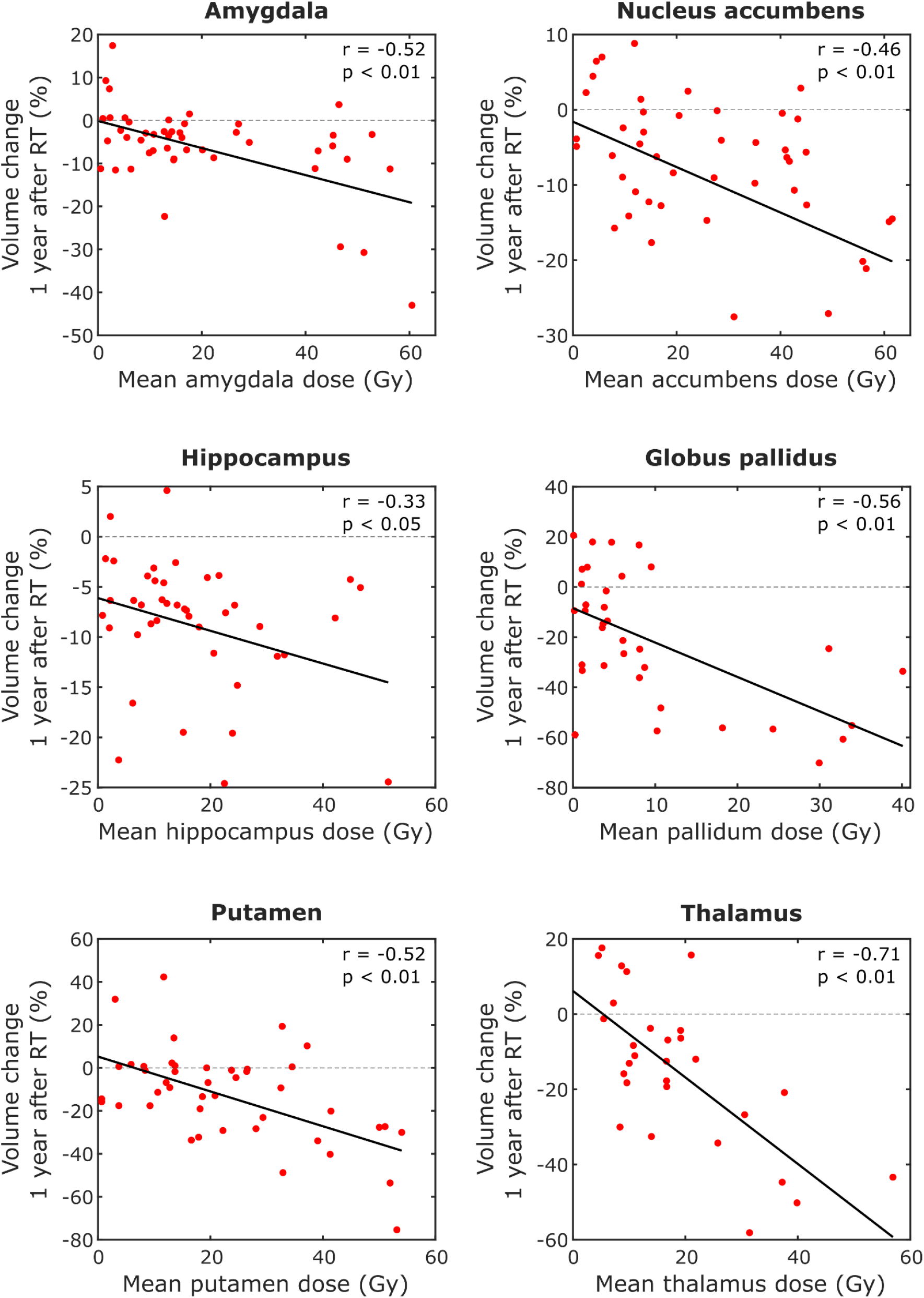
Scatterplots showing the relation between mean RT dose and volume loss in the structures where this was significant, with fitted linear regression lines.

The sensitivity analysis done to assess the effect of chemotherapy on this relation is shown in **Supplementary Table 1**. It did not result in a change in direction or effect size of the results, and chemotherapy administration did not significantly affect GM volume.

### Hippocampal volume nomograms

In this cohort 22 patients were within the age range of 52 to 72, and thus were entered into the nomograms from the UK Biobank, which are shown in **Figure 4**. All patients show an increase in hippocampal age, with a median increase of eleven years (range 2 - 20 years). Accordingly, the percentile within the nomogram dropped for all patients, meaning that their hippocampal volume shows a decrease compared to their peers of the same age.

**Figure 4.**
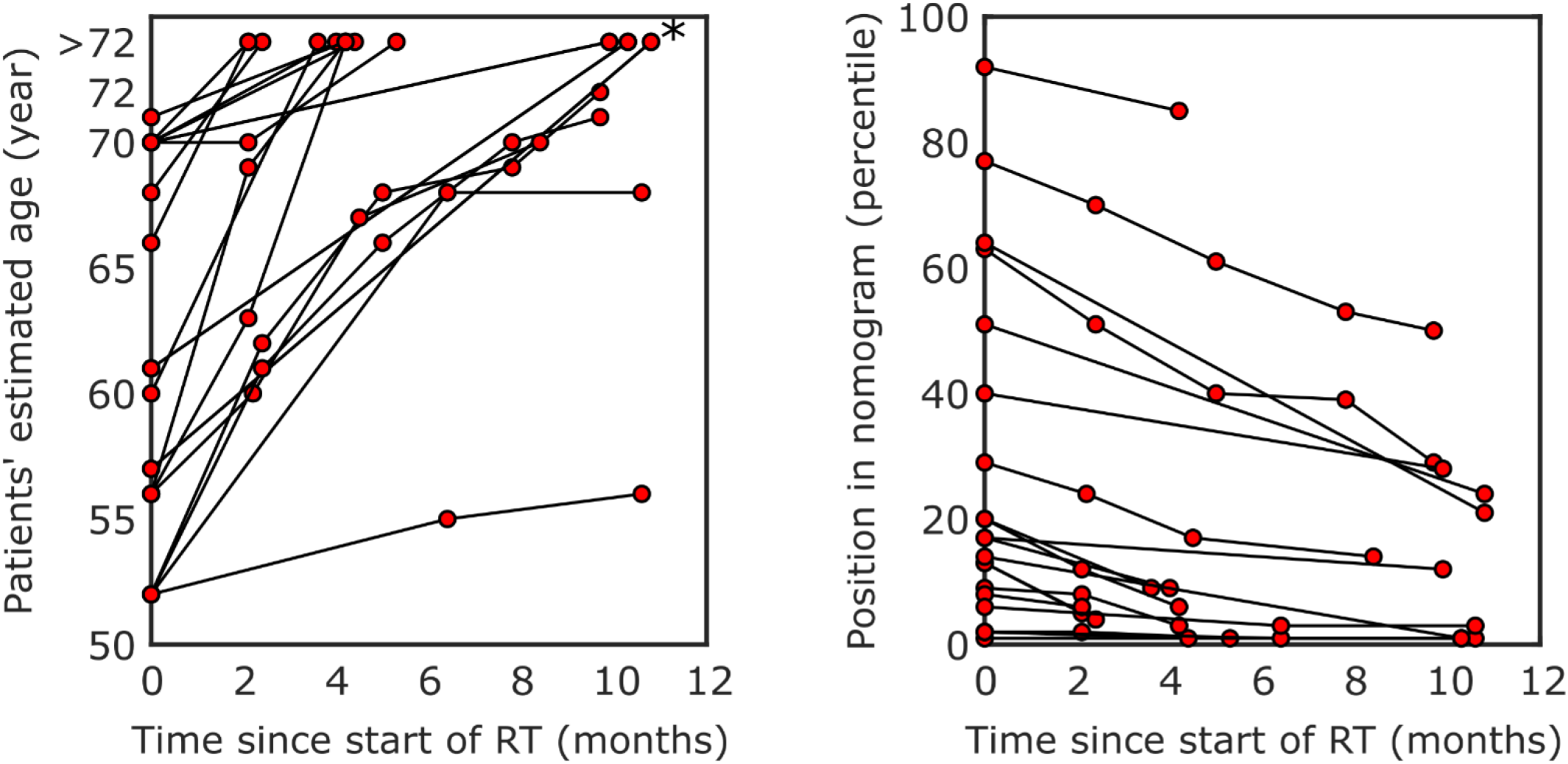
Change in patients’ hippocampal age (left) and position within the nomogram (right) based on the UK Biobank [24].*Age within the nomogram has a maximum of 72.

## Discussion

We have found that all subcortical deep grey matter structures, with the exception of caudate nucleus, show dose-dependent volume loss after RT. For the hippocampus, we have also shown that, based on data from the normal population, its volume-based age increases with up to twenty years during the year post-radiation.

The amygdala and hippocampus have shown to be susceptible to radiation damage in previous studies. Seibert et al. [8] studied MRIs before and one year after RT of 52 patients with primary brain tumours. Automatic segmentation of the hippocampus was followed by relating the difference in volume to the mean dose received. They found a significant correlation with a Pearson correlation coefficient of −0.24. Furthermore, they found that the hippocampus showed significant volume loss after high-dose RT (defined as >40 Gy). This in contrast to low-dose (<10 Gy), which showed no significant relation with post-RT volume. A linear mixed-effects model resulted in a volume loss rate of 0.13%/Gy, which is similar to our observed loss rate of 0.16%/Gy.

The volumetric changes in the amygdala after RT have been studied by Huynh-Le et al. [9] in the same cohort of 52 patients. A significant Pearson correlation of −0.28 was found for amygdala volume and mean dose, with a volume loss rate of 0.17%/Gy. The difference to our findings of a correlation coefficient of −0.52 and volume loss rate of 0.30%/Gy could be explained by the difference in censoring method. They censored amygdalae manually when a visual inspection deemed the segmentation to be poor, whereas we censored more strictly by censoring any organ within the PTV. This meant that they have more data points within higher dose regions, which could have led to a different slope and correlation coefficient.

A link between the volumes of these structures and cognitive outcomes has been thoroughly examined in other brain diseases. Particularly In Alzheimer’s disease, available evidence points towards a strong relation between subcortical GM structures and cognitive impairments for each of the structures we studied except for globus pallidus [14–16]. Furthermore, cognitive impairments in Parkinson’s disease [26,27], multiple sclerosis [28], Huntington disease [17], as well as in normal ageing [12], have been linked with the volume of at least one of the subcortical GM structures. **Supplementary Table 2** gives an overview of some available literature per GM structure.

The effect of hippocampal dose and neurocognitive outcomes was first shown by Gondi et al. [10]. They showed that radiation dose of >7.3 Gy to 40% of the bilateral hippocampi was associated with an impairment in Wechsler Memory Scale-III Word List delayed recall. The same group conducted phase II and phase III trials, the latter studying the effect of whole brain radiotherapy (WBRT) with or without hippocampal avoidance [29,30]. They found hippocampal avoidance WBRT, in combination with the N-methyl-D-aspartate inhibitor memantine, preserves cognitive function while maintaining the same overall and progression-free survival.

In this study the investigation of caudate nucleus volume change in relation to the local dose was inconclusive. One explanation could be that the quality of the segmentation is region dependent. While generally the segmentations of SPM/CAT12 is highly reproducible [31–33], among the investigated regions caudate requires the largest sample size to achieve the same statistical power as compared to other regions [34]. The caudate nucleus shares a relatively large interface with the ventricles (**Figure 2**), making it highly susceptible to partial voluming artefacts, which may lead to errors in segmentation.

Our results challenge us to reconsider the currently used sparing strategies in radiation treatment of brain tumours. Presently, hippocampal sparing RT has been adopted in several institutions. However, sparing the dose in the hippocampus leads to higher doses in surrounding cerebral tissues, which we have shown to be susceptible to radiation-induced damage as well [35]. Future research has to focus on the relation between clinical outcomes (including cognitive and motor function) and morphologic changes, both in the entire brain and in selected structures. This way we can conclusively say which structures should be avoided in RT planning to prevent radiation-induced damage. Specific sparing of healthy brain is possible with novel techniques such as proton therapy and VMAT. This could lead to improved cognition and quality of life in patients undergoing treatment for brain tumours.

There are several limitations to this study. Firstly, we have a relatively limited sample size due to the strenuous inclusion criteria. However, these criteria ensure that the quality of the imaging used in analysis are optimal, meaning more reliable and replicable results. The censoring of the PTV also means exclusion of several subcortical GM structures, but this again is to ensure reliable automated measurements. Attempts could have been made to manually delineate these structures, but this would have added an extra variable to the dose/volume relation (manual vs automatic segmentation in high and lower dose, respectively). Additionally, it is unclear which method gives the most reliable results in patients who underwent RT. In previous works [36–38], the automated segmentation method used in the current study was rigorously compared to manual segmentation of subjects’ T1 MRI data as well as brain phantoms representing a wide range of settings (noise, artefacts, etc.). It was found that CAT12 performed on a comparable level versus manual segmentation in healthy subjects as well as patients with ischemic stroke or temporal lobe epilepsy, suggesting it is reliable for segmentation in RT patients. Another consideration is that susceptibility to radiation-induced volume loss of brain tissue might differ between patients. As each patient provides multiple organs for examination, this could have impacted the found results. We could not correct for this in the current study, as our sample size limited our ability to apply multilevel modelling of the dose/volume relation.

Secondly, the patients in our cohort did not only undergo RT. Many also received chemotherapy, which has been linked to cerebral changes in non-neurological malignancies [39,40]. Our analysis focussed on the association between RT dose and volume, and by relating these two factors to each other we have limited the effect of chemotherapy as much as possible. Furthermore, a sensitivity analysis including chemotherapy in the model did not give different results, suggesting its role is limited.

Finally, the absence of neurocognitive outcome data from this cohort means we cannot yet give clinical recommendations on which organs to spare.

In conclusion, subcortical grey matter structures show susceptibility to dose-dependent volume loss after radiotherapy. If neurocognitive outcomes are related to this phenomenon, current RT strategies need to be revised, in order to improve patients’ quality of life after cancer treatment.

## Data Availability

No data is available

## Abbreviations

CAT12: Computational Anatomy Toolbox 12
CT: computed tomography
FWER: Family-wise error rate
GM: Grey matter
MRI: Magnetic resonance imaging
PALM: Permutation analysis of linear models
PTV: Planning target volume
RT: Radiotherapy
SPM: Statistical Parametric Mapping
TFE: turbo fast echo
WBRT: Whole-brain radiotherapy

